# Novel Spatiotemporal Feature Extraction Parallel Deep Neural Network for Forecasting Confirmed Cases of Coronavirus Disease 2019

**DOI:** 10.1101/2020.04.30.20086538

**Authors:** Chiou-Jye Huang, Yamin Shen, Ping-Huan Kuo, Yung-Hsiang Chen

**Affiliations:** School of Electrical Engineering and Automation, Jiangxi University of Science and Technology, Ganzhou, Jiangxi, 341000, China; Computer and Intelligent Robot Program for Bachelor Degree, National Pingtung University, Pingtung 90004, Taiwan; Department of Mechanical Engineering, National Pingtung University of Science and Technology, Pingtung 91201, Taiwan

**Keywords:** Confirmed cases forecasting, COVID-19Net, parallel deep neural network

## Abstract

The coronavirus disease 2019 pandemic continues as of March 26 and spread to Europe on approximately February 24. A report from April 29 revealed 1.26 million confirmed cases and 125 928 deaths in Europe. This study proposed a novel deep neural network framework, COVID-19Net, which parallelly combines a convolutional neural network (CNN) and bidirectional gated recurrent units (GRUs). Three European countries with severe outbreaks were studied—Germany, Italy, and Spain—to extract spatiotemporal feature and predict the number of confirmed cases. The prediction results acquired from COVID-19Net were compared to those obtained using a CNN, GRU, and CNN-GRU. The mean absolute error, mean absolute percentage error, and root mean square error, which are commonly used model assessment indices, were used to compare the accuracy of the models. The results verified that COVID-19Net was notably more accurate than the other models. The mean absolute percentage error generated by COVID-19Net was 1.447 for Germany, 1.801 for Italy, and 2.828 for Spain, which were considerably lower than those of the other models. This indicated that the proposed framework can accurately predict the accumulated number of confirmed cases in the three countries and serve as a crucial reference for devising public health strategies.

## I. INTRODUCTION

CORONAVIRUS disease 2019 (COVID-19) is caused by a highly contagious novel virus and was first discovered in Wuhan City in Hubei, China, where the pandemic initially began. The pandemic quickly spread across China and then the entire world. Because of the high contagion rate of COVID-19 and the likelihood of an outbreak, a lockdown was imposed on Wuhan City, which has a population of nearly 11 million people. Meanwhile, the Chinese government implemented a series of strict measures to control the pandemic. Fig. 1 31 illustrates the timeline of the COVID-19 pandemic.

**Figure 1.**
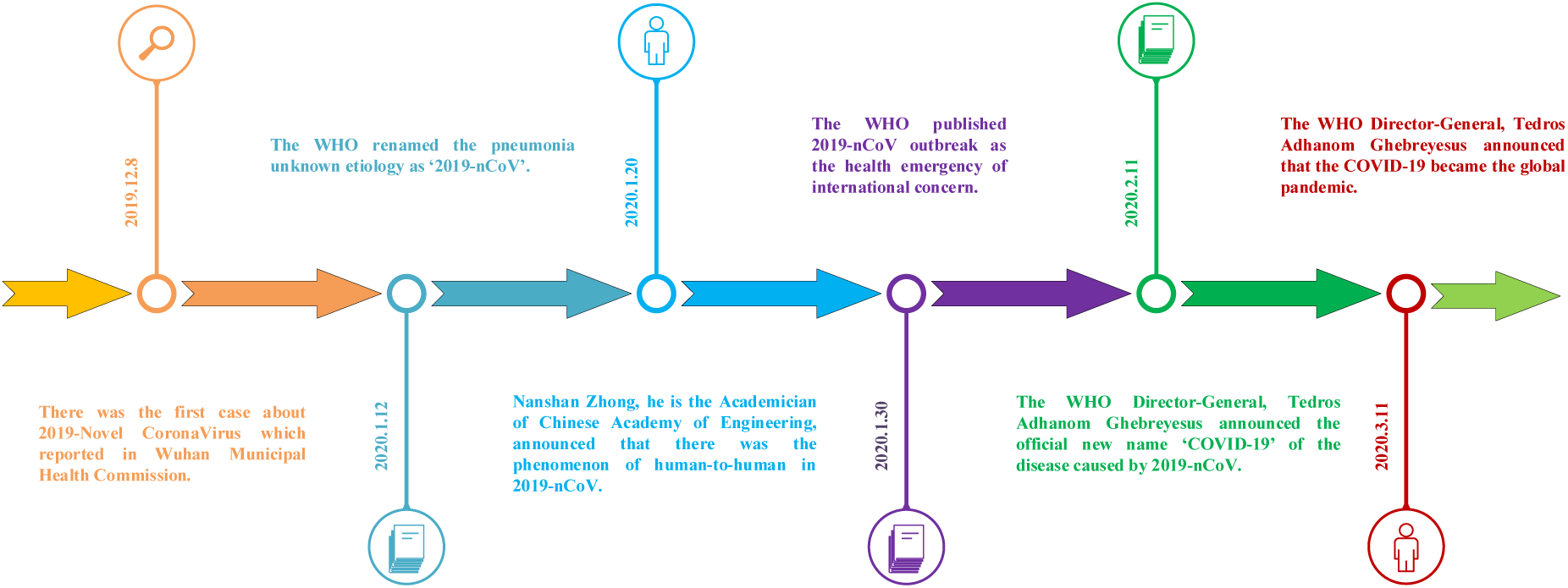
Timeline of the COVID-19 pandemic

In December 2019, a pneumonia case of unknown cause was discovered. Shortly afterward, patients with similar symptoms appeared across China and substantially burdened the healthcare system. On January 12, 2020, the World Health Organization (WHO) named the pneumonia 2019-nCoV. The disease was then confirmed to be contagious between people by Zhong Nanshan, an academician of the Chinese Academy of Engineering, on January 20, 2020. Ten days later (January 30), the WHO designated 2019-nCoV a public health emergency of international concern. On February 11, 2020, WHO Director-General Tedros Adhanom Ghebreyesus officially renamed the disease COVID-19 during an international meeting in Geneva. On March 11, 2020, Tedros announced a global outbreak of COVID-19. Mahase [1] reported in the BMJ that UK Prime Minister Boris Johnson warned citizens to avoid contact with people. Imperial College London indicated that COVID-19 is highly contagious to older adults aged over 70 years and pregnant women.

As of 11:00 am Beijing Time on March 16, the accumulated number of confirmed cases in Italy has reached 24 938, with 20,794 active cases, 1518 severe cases, 2335 fully recovered cases, and 1809 deaths. Italy (60.43 million people) has only a 20% larger total population than does South Korea (51.63 million people). However, the number of confirmed cases in Italy is three times of that South Korea. The number of deaths presents the greatest difference; that of Italy (1809) is 24 times that of Korea (75). Overall, the COVID-19 mortality rate in Italy exceeds 7%, which is substantially higher than the global average mortality rate (3.7%) and is the highest rate worldwide. Italy implemented measures against the pandemic earlier than did other countries. The day after COVID-19 was designated a public health emergency of international concern by the WHO, the Italian government became the first among European Union countries to announce a national health emergency and ceased all flights to and from China. On February 22, 2020, Italy imposed lockdowns on multiple cities, and the military was deployed to assist with pandemic prevention 3 days later. Recently, Italy has deployed healthcare personnel across the country to contain the pandemic. Italy’s severe situation and its worldwide leading mortality rate may be attributable to the fast spread of the virus and numerous citizens’ unwillingness to reduce social activities despite warnings from the government. Meanwhile, an increasing number of countries have implemented strict pandemic prevention measures. After Italy, Spain imposed multiple strict measures in which the central government announced a state of emergency for 14 days, during which all people were requested to remain indoors unless commuting to work or purchasing daily necessities. In addition to schools, which were closed previously, all shops and commercial areas unrelated to sustaining daily necessities were required to cease operation. On March 14, 2020, a lockdown was imposed on Madrid. Madrid, Spain on Saturday, March 14 became the second European country to impose a nationwide lockdown, ordering its 47 million people to mostly stay in their homes as coronavirus cases surge and the government announced that the wife of the Spanish prime minister had tested positive for the virus [2]. Staring from the next day, the police began to use unmanned drones for patrols and ask crowds in parks to leave and remain at home. However, the pandemic in Spain has not subsided. The accumulated number of confirmed cases in Spain has reached 74 386, and this number is higher than that of the five most affected Chinese provinces excluding Hubei combined (i.e., Guangdong, Henan, Zhejiang, Hunan, and Anhui), despite the total population of Spain being 12% of the combined population of these five provinces. The incremental rate of confirmed cases in Spain is also greatly concerning. The accumulated number of confirmed cases was 3146 on March 13, 5232 on March 14, and 7753 on March 15, which presents a daily incremental rate of 50%. In addition, the number deaths in Spain reached 288 and a 3.7% mortality rate, which is the highest after that of Italy.

On February 9, 2020, Fernando Simón, the Director of the Center for Coordination of Health Alerts and Emergencies in Spain, stated that the country would only have a few confirmed cases. However, a report by the center on March 6 warned that the number of deaths in nearby Italy was increasing rapidly and that the pandemic may spread to Spain. At that time, the number of confirmed cases in Spain was roughly 280, with four deaths. Two weeks later, the number of deaths in Spain exceeded 7000, three times that disclosed by Iran. The statistics reported by the Spanish government on March 30 indicated that 813 people died because of COVID-19 in the past 24 h, which caused the number of deaths in Spain to reach 7340, during which the number of confirmed cases increased by 6398. Consequently, the number of accumulated confirmed cases reached 85 195 in Spain, the third highest number worldwide. The pandemic in Spain appeared to have subsided in the past day. However, a sudden increase in the number of confirmed cases and deaths forced the Spanish government to impose further lockdown measures: Since March 30, all people have been required to remain at home for at least 2 weeks, except for those engaging in essential occupations.

On March 11, 2020, the Chancellor of Germany, Angela Merkel, hosted a press conference jointly with Jens Spahn, the Federal Minister of Health, and Lothar Wieler, the President of the Robert Koch Institute. For the first time, Merkel publicly addressed her opinion of COVID-19 and stated that the public is currently not immune to this disease. Merkel warned that because no vaccine or medication has been developed for COVID-19, 60%–70% of the German population is expected to be infected at the current rate according to expert analysis. A few hours after the press conference, the WHO characterized COVID-19 as a pandemic, when the accumulated number of confirmed cases in Germany reached 2000, with four cases of death. This marks the zero-death record held by Germany while the pandemic spreads across Europe.

In the past week, the number of confirmed cases in Germany has increased considerably, with daily increases of over 1000 cases. To contain the pandemic, the German government closed its borders with France, Switzerland, and Austria at 08:00 on March 16. The German railway also progressively reduced the number of domestic trains in service. Social activities in each German state are being gradually reduced. Events with more than 50 participants must be canceled; schools, bars, and nightclubs have been closed; some companies and governmental units allow their employees to work from home; and restrictions were imposed on foreign travel. In response to COVID-19, the German government has conducted research in the Robert Koch Institute since January 6, and the scale of the research continues to increase. In addition to its comprehensive laboratory system and highly trained personnel, Germany began conducting screening tests early to identify patients in advance. After the contagion rate and number of patients are reduced, the national healthcare system can allocate more capacity to provide treatment for all patients instead of passively conducting screening tests after patients experience severe symptoms and require hospitalization. Germany currently has the lowest COVID-19 mortality rate among European countries with populations of more than 10 million people. According to 2017 Organization for Economic Cooperation and Development statistics, the number of ward beds per person in Germany was the highest among European countries, and an average of 8 beds are available for every 1000 people. A crucial factor in containing pandemics is the density of intensive care units (ICUs). Patients with severe COVID-19 symptoms must receive treatment in ICU wards and generally require artificial ventilation. According to the German Federal Ministry of Health, Germany currently has 28 000 intensive care beds, 25 000 of which are equipped with artificial ventilation systems. We assume that Germany’s healthcare system possesses enough facilities to contain the pandemic for the total population. However, many of these beds are already occupied. Consequently, this assumption is valid only if the national healthcare system does not collapse. A key factor in preventing this pandemic is reducing the viral contagion rate and not overwhelming the healthcare system, which can be extremely difficult. The number of intensive care beds in Italy is roughly 5000. If the pandemic continues or becomes more severe, Germany’s large number of available beds will be a notable advantage in containing the pandemic.

As of 16:00 on March 20, the accumulated number of confirmed cases in Germany is 19 356, with 52 deaths and a mortality rate of 0.26%. However, the situation in Germany is not optimistic. Political and academic figures, including Minister Spahn, warned that the pandemic has not peaked. Before the COVID-19 outbreak, the German healthcare system already faced labor shortages. Experts warned that when the real peak of the pandemic arrives, Germany may lack nurses despite having sufficient ventilators. Local health departments lack epidemiology surveyors; hence, during the pandemic peak, whether Germany can continue to effectively track the contact history of cases and stop chains of infection remain questionable. Concerns have emerged regarding European countries with close interactions with Germany, including the United Kingdom and Switzerland. If these countries fail to contain the pandemic, Germany may face continuous streams of imported cases because the strict border controls currently imposed in Europe cannot continue indefinitely.

In response to COVID-19, which is considered a “never before threat” to Germany, the German government clearly stated that additional restrictions may be imposed on the daily life of citizens. Merkel also emphasized that the government is ready to take any necessary measures to help the country overcome the pandemic. As the country with the largest population, economic activity, and number of European Parliament seats in the European Union, Germany is a critical reference for the global community to assess the progression of COVID-19. Currently, the pandemic in China appears to have subsided considerably, while outbreaks continue in other countries, particularly European countries such as Italy, Spain, and Germany. On March 19, 2020, the number of newly confirmed cases in Italy surpassed 4000, and that in Spain and Germany each surpassed 2000. The COVID-19 outbreak substantially threatens the global health system and considerably affects the global economy. Decreases in European and US stock markets have led to notable economic losses. Extracting information from the spatiotemporal data of the pandemic is increasingly crucial to helping countries determine next-stage pandemic prevention measures and planning resource allocation and aid among cities and countries.

The remaining sections of this paper are structured as follows. Section 2 presents a literature review on articles recently published by relevant scholars. Section 3 details the model proposed in this study and the principles behind models. Section 4 discusses the analysis results, and Section 5 presents its conclusions.

## II. RELATED WORKS

Commonly used COVID-19 prediction models are divided into two types: those using mathematics or artificial intelligence (AI) black box algorithms to construct models and analyze the progression and contagion of the pandemic. Li et al. [3] used a mathematical approach to construct a function model and describe the daily number of newly confirmed cases and deaths in Hubei. They used nonlinear regression to obtain essential parameter values, summarize the pandemic trend in Hubei, and predict that the pandemic will end in Hubei on March 10. Nishiura et al. [4] estimated the serial intervals of COVID-19 patients. Using a truncated dataset with days as the unit, they employed a double-interval Bayes likelihood function to estimate serial intervals. Under a 95% confidence interval, their results revealed a mean of 3.9 d between successive cases in a chain of transmission. Tang et al. [5] used a temporal dynamic model to simulate COVID-19 transmission in China. Markov chain Monte Carlo methods were used to fit the pandemic data of the country, and Geweke’ s diagnostic was used to assess model convergence. Their experiment suggested that 58%–76% of contagion sources must be isolated to contain the pandemic. Fan et al. [6] used data on the population distribution and flow of Wuhan City to predict COVID-19 cases imported from nearby cities and provinces. Their results revealed that the population of other cities or provinces was highly correlated with the daily number of confirmed cases in Wuhan. Roosa et al. [7] predicted pandemic progression by using three mathematical models: a generalized logistic growth model, the Richards growth model, and a sub - epidemic wave model. Their results verified that the sub - epidemic model most accurately predicted the pandemic situation in Hubei on February 15. Zhong et al. [8] used the susceptible–infected–removed model and epidemic data combined with parameter estimation to estimate the accumulated number of confirmed cases in China. They predicted that the COVID-19 outbreak in China will gradually subside between early May and late June.

The Diamond Princess cruise is considered a major epidemic zone of COVID-19 and has received considerable attention from the global community. Nishiura [9] used the infection timeline of the Diamond Princess to back-calculate the incidence rate under a 95% confidence interval and revealed that implementing isolation measures on February 5 reduced the number of confirmed cases to 776. Currently, Internet and big data platforms are gradually improving to incorporate comprehensive functions. This indicates the feasibly of using AI to resolve disease-related problems [10]. For example, in response to the increasing need for resources during the severe acute respiratory syndrome (SARS) outbreak from China in 2002, Yu et al. [11] used a multitarget and multicycle mixed-integer programming model to determine the optimal allocation and provision of temporary facilities and resources. They used Language for Interactive General Optimizer to optimize facility site selection and suggested that selecting the sites of temporary medical facilities built in late February substantially affects subsequent pandemic prevention. McAleer [12] employed the Global Health Security Index to assess 195 countries worldwide and compared COVID-19 with SARS and Middle East respiratory syndrome. The results indicated that the global community could better contain COVID-19 than the other two diseases, with the index score increasing to 51.9.

For deep learning, Metsky et al. [13] employed the CRISPR-Cas13 system to compare COVID-19 virus nucleic acid sequences with those of other representative viruses. They also tested various COVID-19 screening methods and identified ADAPT as the most accurate. Guo et al. [14] used a bi-path convolutional neural network (CNN) to construct a virus host prediction model. They used the basic local alignment search tool to compare the gene sequences of two virus hosts and determined that the area under curve approach most accurately described human hosts. Yang et al. [15] combined a long short-term memory (LSTM) model and the Susceptible-Exposed-Infected-Resistant (SEIR) model and used the population flow data obtained near January 23, 2020 and COVID-19 epidemic parameters to predict the pandemic peak in China. Comparing the prediction curves derived using these two models and that plotted using actual data indicated that the number of infected patients would reach a peak of 4000 between February 4 and February 7. Anastassopoulou et al. [16] used the susceptible–infectious–recovered–dead model to examine the medical data of Hubei acquired between January 11 and February 10, 2020 and predict mortality and recovery rates through 95% confidence intervals [17]. Their results showed that the accumulated number of confirmed cases could reach 45 000–180 000 on February 29, and the number of deaths could exceed 2700. Kucharski et al. [18] used a random transmission model to predict the number of confirmed cases in Wuhan and imported cases from Wuhan. Their results indicated that viral transmission could subside in Wuhan in late January 2020; however, new outbreaks could occur in regions with transmission potential similar to that of Wuhan. Huang et al.[19] proposed to use the deep neural network CNN model to predict cumulative confirmed cases of COVID-19 in seven regions of Wuhan et al. By comparing the results with those of the MLP, LSTM and GRU models, the overall effect of CNN was found to be better than the other models in that study.

Few scholars have used deep learning to predict pandemic trends because this approach requires much data. To fully utilize available pandemic data, the time-variant characteristics of pandemic progression and the strong spatial correlation between neighboring countries and cities with high transmission potential must be considered. Accordingly, a parallel mixed deep learning network was employed in this study to extract the spatiotemporal features of COVID-19 transmission and predict subsequent pandemic trends.

## III. METHODOLOGY

Data were acquired from the daily situation reports disclosed by the WHO [20] and information in the GitHub website [21]. To ensure the integrity and rigorousness of research procedures, we used data from January 22 to April 28, during which 98 pieces of data were collected daily and divided into 93 samples. The first 83 samples were used as the training set, and the remaining 10 samples were used as the testing set. Data accumulated over a 5-d period were used to predict the accumulated number of confirmed cases of the subsequent day. The data were from three European countries with severe situations: Germany, Italy, and Spain. The data were divided into two parts:

1. The first part consisted of six characteristic factors from each country: the daily number of newly confirmed cases, deaths, and recovered cases and the accumulated number of confirmed cases, deaths, and recovered cases.
2. The second part consisted of three characteristic factors—the accumulated number of confirmed cases from the three countries.

Fig. 2 illustrates the geographical locations of the three countries.

**Figure 2.**
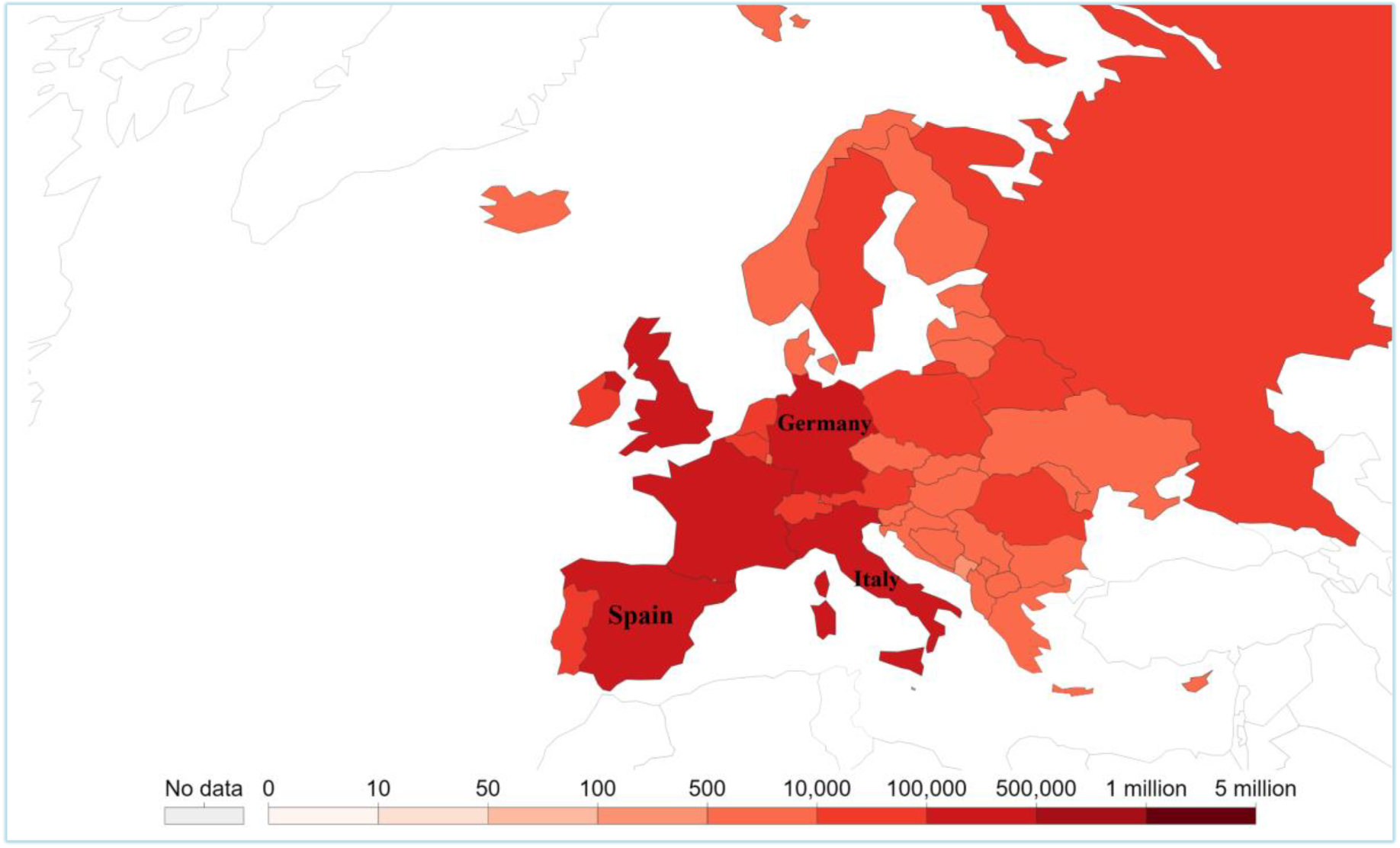
Geographical locations of Germany, Italy, and Spain. [22]

### A. Convolutional Neural Network

CNNs are a typical class of deep neural networks, [23, 24] and have strong feature-learning ability. A CNN can be divided into layers to extract features from a dataset. Fig. 3 presents the structural diagram of a CNN.

**Figure 3.**
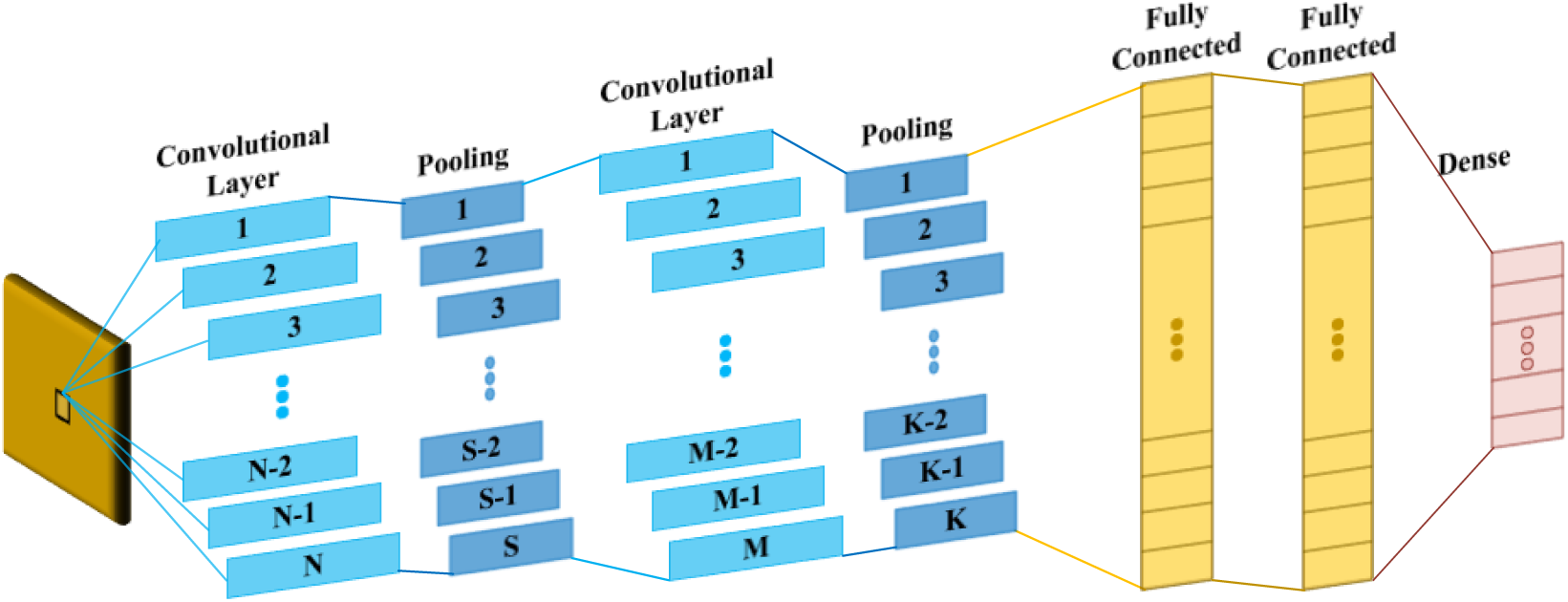
CNN structure.

A CNN mainly comprises convolutional, pooling, and fully connected layers. Convolutional layers are the core structure where most computations occur. When data pass through the kernel, features are extracted and sent to pooling layers, which ensure that features remain invariant. The resulting feature maps are then compressed to reduce computation loads. Finally, fully connected layers transform all features into onedimensional (1D) vectors and send them to the dense layer for outputs.

### B. Bidirectional Long Short-Term Memory Neural Network

A bidirectional LSTM (BiLSTM) model [25], [26] presents an improved framework compared with LSTM [27],[28]. The neuron structure of BiLSTM is similar to that of LSTM, as illustrated in Fig. 4(a) and 4(b). LSTM comprises an input gate, output gate, and forget gate.

**Figure 4.**
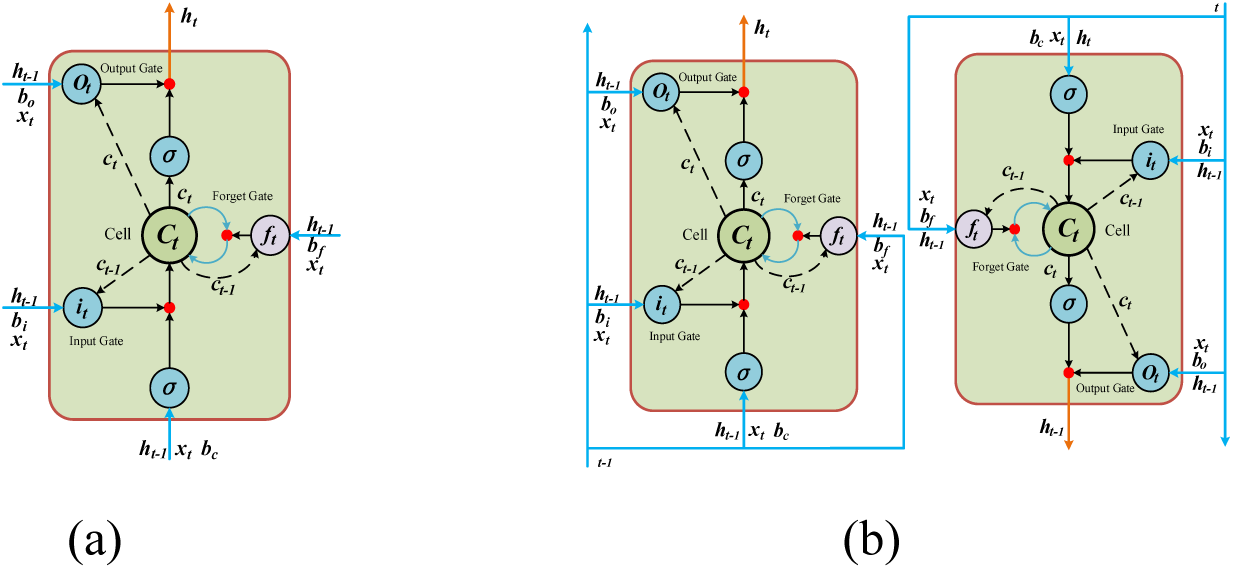
(a) Neuron structure of an LSTM model; (b) neuron structure of a BiLSTM model.

The input gate is expressed using (1), in which current input *x_t_* and previous information and memory (i.e., *h_t_*_−1_ and *c_t_*_−1_) are selectively retained according to corresponding weights and are sent to the forget gate. The corresponding weight matrices are *W_xi_*, *W_hi_*, and *W_ci_*, and *b_i_* denotes the bias matrix of the input gate.

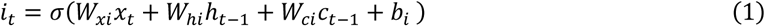

The forget gate is expressed using (2) and (3). In (2), current input *x_t_* and previous information and memory (i.e., *h_t_*_−_*_1_* and *c_t_*_−_*_1_*) are selectively forgotten according to weight matrices *W_xf_*, *W_hf_*, and *W_cf_*, and *b_f_* is the bias matrix of the forget gate. In (3), information *i_t_* retained in the input gate is applied to the tanh function and added to previously forgotten information *f_t_c_t-1_* to derive the target retained information *c_t_*, where *W_xc_* and *W_hc_* are the weight matrices of the forget gate, and *b_c_* is the bias matrix of the forget gate.

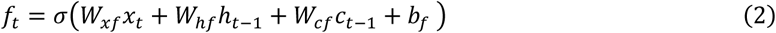

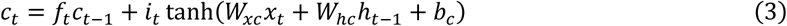

The output gate is acquired by passing current input *x_t_*, previous information *h_t_*_−1_, and currently retained memory *c_t_* through the sigmoid layer. Then, *c_t_* is standardized using tanh and multiplied to the output of the sigmoid layer to derive output information *h_t_*. In (4) and (5), *W_xo_*, *W_ho_*, and, *W_co_* are the weight matrices of the output gate, and *b_0_* is the bias matrix of the output gate.

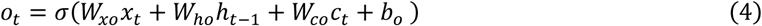

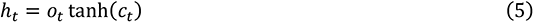

A BiLSTM model is an LSTM model with backpropagation learning, as presented in Fig. 4(b).

### C. Bidirectional Gated Recurrent Unit

A gated recurrent unit (GRU) [29],[30] resolves the vanishing gradient problem in a recurrent neural network and uses an update gate and reset gate, as displayed in Fig. 5(a). These two gates control output information, retain previous information, and require fewer parameters than does an LSTM model. Accordingly, a GRU-based neural network is more efficient than an LSTM model.

**Figure 5.**
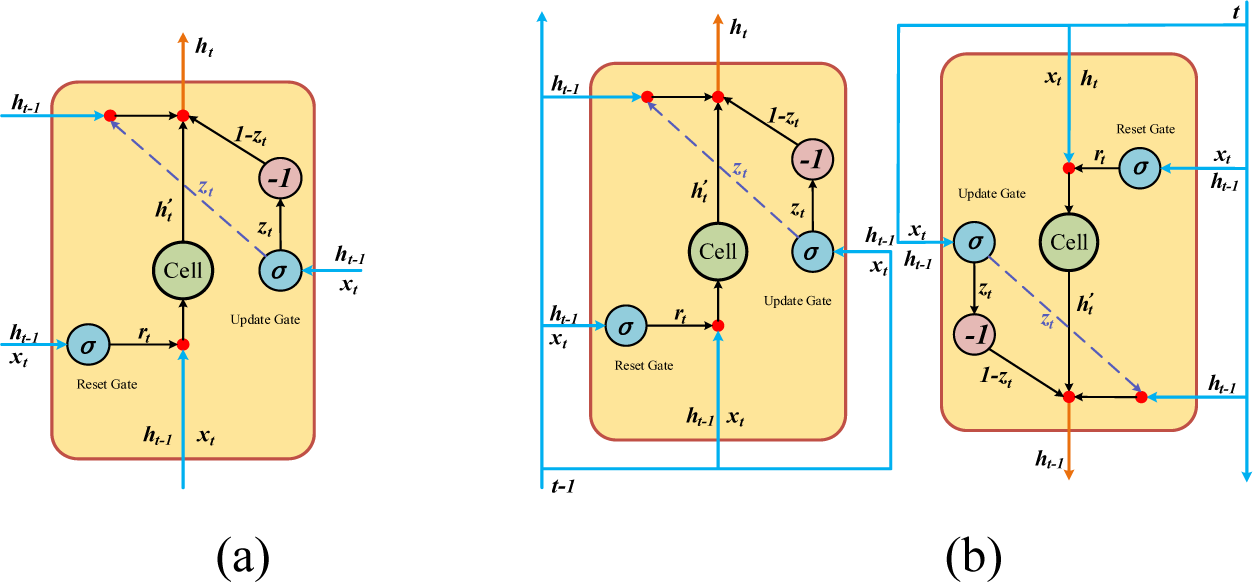
(a) GRU neuron structure; (b) BiGRU neuron structure.

The update gate controls information sent previously. In (6), the update gate can selectively retain previous information *h_t_*_−1_, and *W*^(^*^z^*^)^ and *U*^(^*^z^*^)^ are the weight matrices of the update gate.

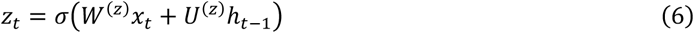

The reset gate selectively forgets previous information *h_t_*_−1_. The computation of (7) is identical to that of (6), and *W*^(^*^r^*^)^ and *U*^(^*^r^*^)^ are the weight matrices of the reset gate.

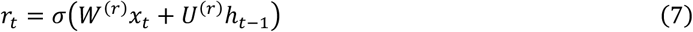

Reset gate output *r_t_* is subjected to matrix multiplication with *Uh_t_*_−1_, as expressed in (8).

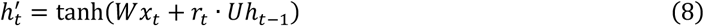

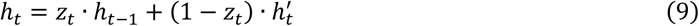

A bidirectional GRU (BiGRU) [31] is similar to a BiLSTM [25] model in which backpropagation learning is introduced on the basis of GRU, as shown in Fig. 5(b).

### D. Proposed Method

Fig. 6 illustrates the flow chart of the proposed algorithm COVID-19Net.

**Figure 6.**
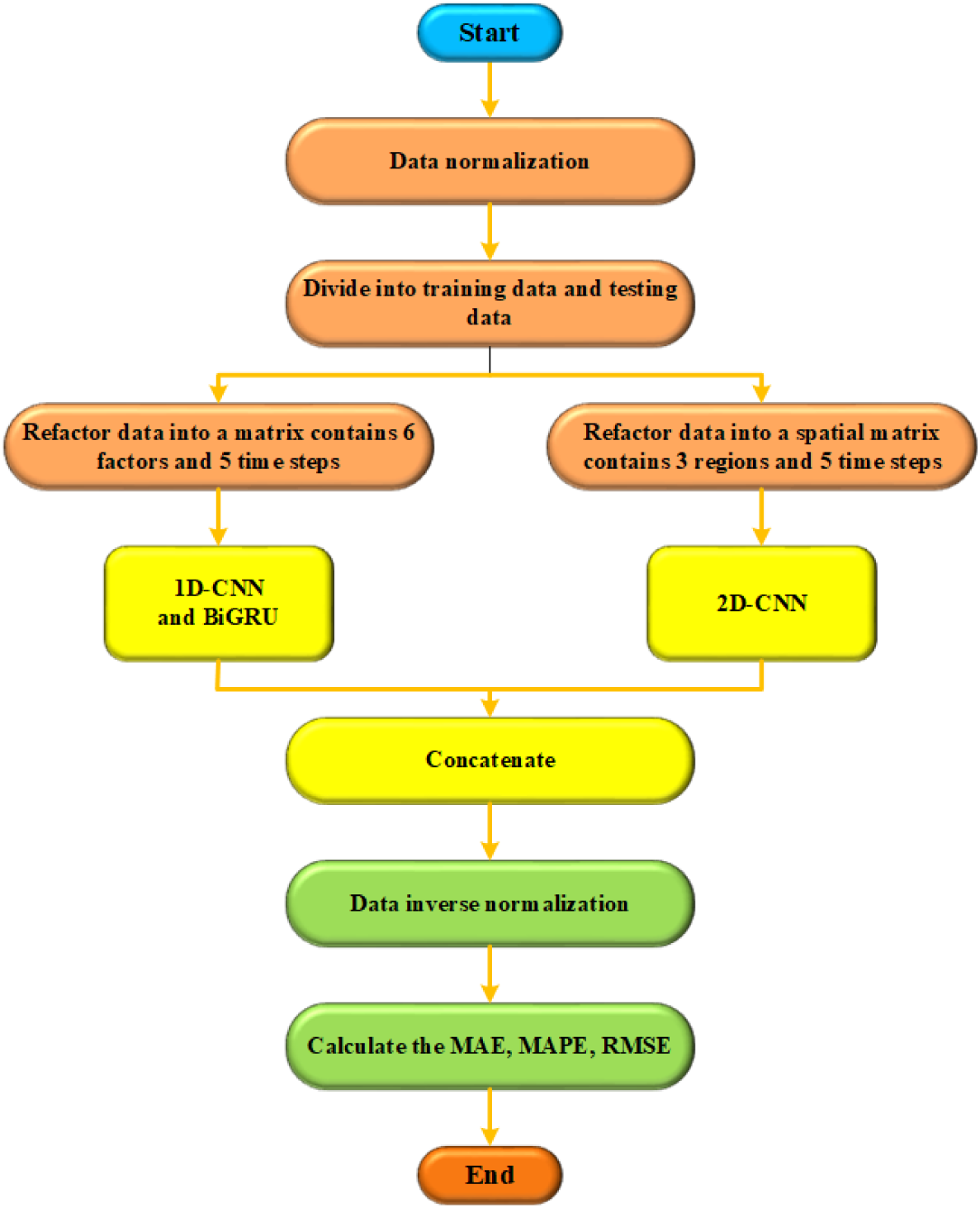
Flow chart of COVID-19Net.

Step 1: Required data are selected from the aforementioned data sources to perform correlation analysis and identify highly correlated data.

Step 2: All data are divided into testing and training sets according to the data collection time.

Step 3: Data are refactored into a matrix with six factors and five time steps named Input 1. Data are refactored into a matrix with three regions and five time steps named Input 2.

Step 4: Because Input 1 is mainly used to extract temporal features, a 1D-CNN and BiGRU-based parallel deep learning network is employed. Because Input 2 is mainly used to extract spatiotemporal features and each country has a specific geographical location and order, a two-dimensional (2D) CNN is employed. The training set refines the models.

Step 5: The testing set is used to test the models and calculate the mean absolute error (MAE), mean absolute percentage error (MAPE), and root mean square error (RMSE).

The COVID-19Net algorithm proposed in this study is parallelly connected using a 1D-CNN [32], a 2D-CNN [33], and BiGRUs to form a mixed deep learning network. Fig. 7 illustrates this framework.

**Figure 7.**
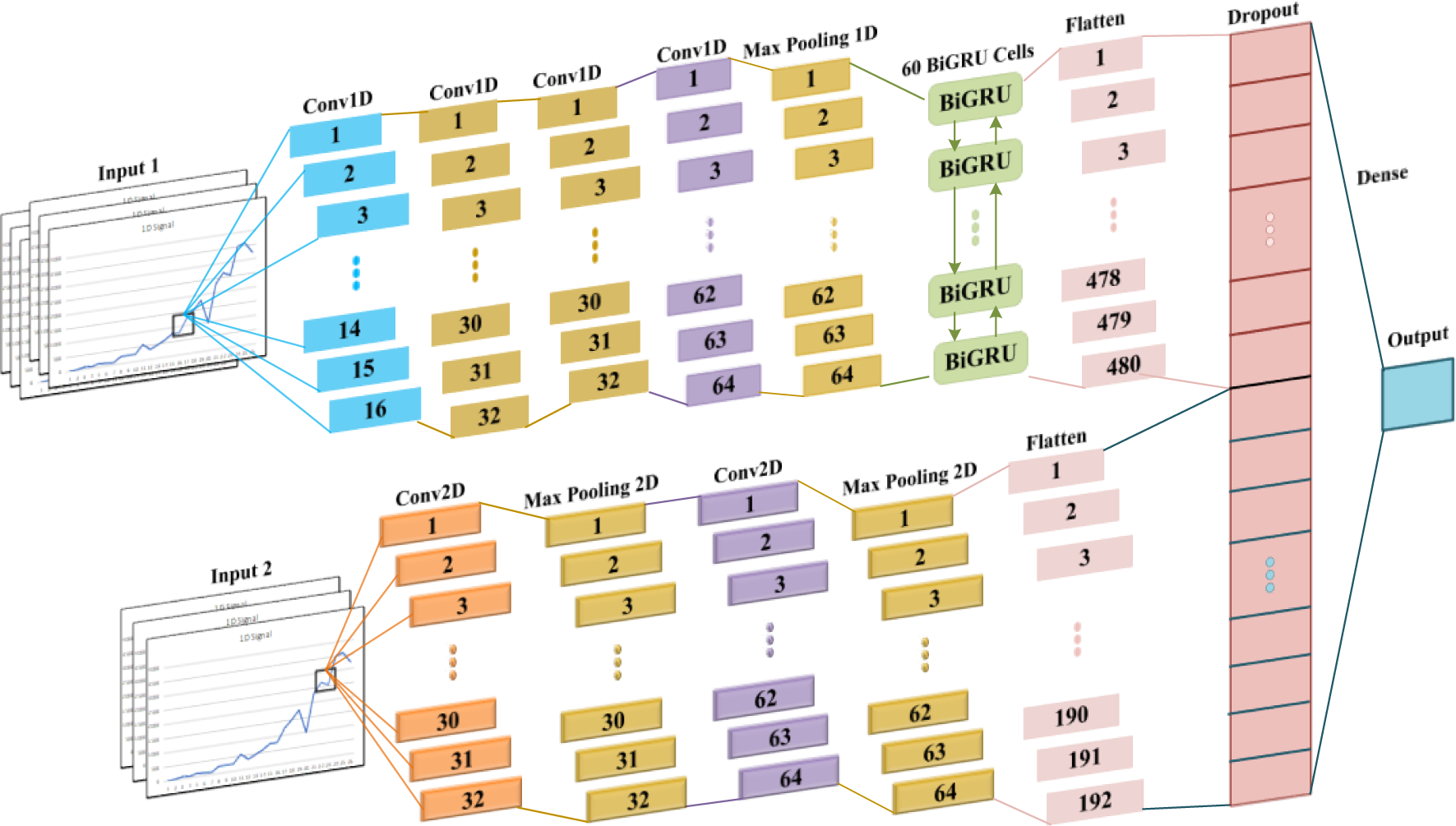
Framework of COVID-19Net.

COVID-19Net was used to process spatiotemporal and temporal data separately. Because Input 1 contained data from each country regarding the daily number of newly confirmed cases, deaths, and recovered cases and the accumulated number of these cases in the past 5 days, temporal features related to the accumulated number of confirmed cases were extracted from these factors. Given the outstanding performance of BiGRUs in learning from time series data, we serially combined a 1D-CNN with BiGRU to extract temporal features from Input 1. The highly contagious nature of COVID-19 and frequent population flows among European countries may result in a strong spatial correlation between their pandemic trends. Therefore, extracting spatial features considerably increased the accuracy of the prediction model. This indicated that features related to changes in the accumulated number of confirmed cases in each country are highly crucial. Based on the parallelly combined 1D-CNN and BiGRUs used for extracting temporal features from Input 1, a 2D-CNN was constructed to extract the spatiotemporal features of the three countries from Input 2 (Fig. 6). The 1D-CNN had 16, 32, 32, 64 convolutional kernels, respectively, each with a kernel size of 3. The size of the corresponding maximum pooling layer was 2. In the 2D-CNN model, the two convolutional layers had 64 and 96 convolution kernels, respectively, and the sizes of the convolution kernels were 3×3 and 4×1, respectively. Because the COVID-19 data employed in this study were insufficient, a dropout layer was used to prevent overfitting.

The COVID-19Net algorithm proposed in this study parallelly combines CNNs and BiGRUs to concurrently process spatiotemporal and spatial features. The proposed algorithm can extract temporal and spatiotemporal features, which provides an alternative for researching data with both temporal and spatiotemporal correlation.

## IV. EXPERIMENTAL RESULTS AND DISCUSSION

The MAE, MAPE, and RMSE equations used in this study are expressed as follows:

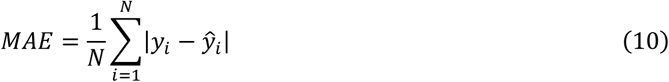

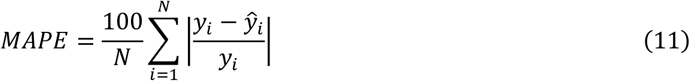

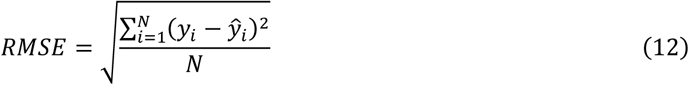

Experimental data were collected from Germany, Italy, and Spain regarding six factors: the daily number of newly confirmed cases, deaths, and recovered cases and the accumulated number of confirmed cases, deaths, and recovered cases. Fig. 8 presents the correlation heatmaps of these factors. The results revealed that the accumulated number of confirmed cases was strongly correlated with other factors in Italy, with all correlation coefficients greater than 0.4 (Fig. 8[b]). Fig. 8(a) and 8(c) displays the correlation heatmaps of Germany and Spain, respectively. The pandemic situations in these countries were less severe than that of Italy. Therefore, the correlation between accumulated confirmed cases and daily recovered cases was low, with correlation coefficients of 0.78 and 0.65. However, the correlation between the remaining factors remained high, with correlation coefficients exceeding 0.9. The results indicated that using these six factors for model training was necessary.

**Figure 8.**
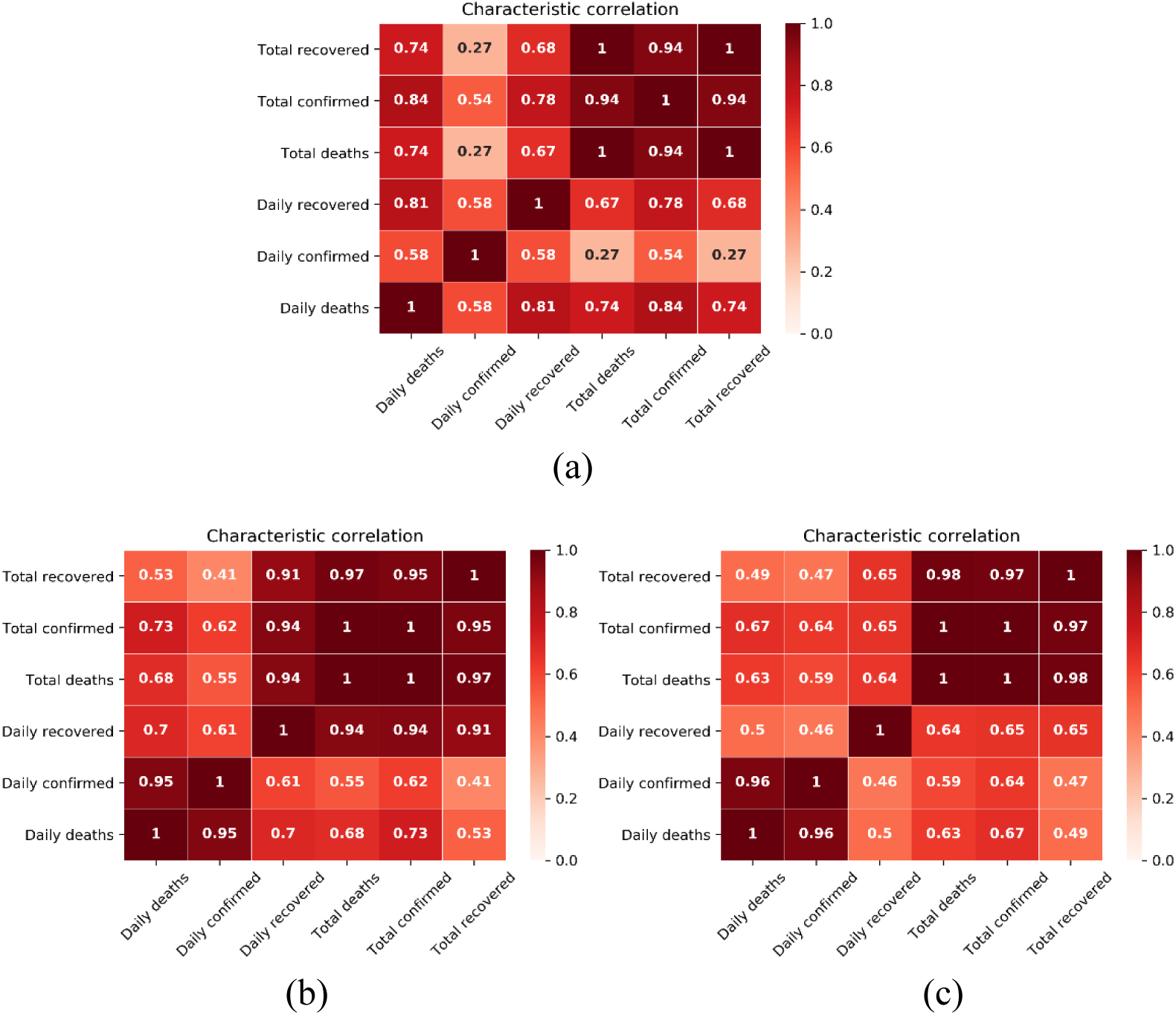
Correlation heatmaps of (a) Germany, (b) Italy, and (c) Spain.

A separate correlation heatmap was created for the accumulated number of confirmed cases in the three countries (Fig. 9). The results indicated correlation coefficients greater than 0.99. This may have been attributable to the geographical proximity of these countries and their frequent interaction, which lead to faster disease transmission. This suggested that extracting spatiotemporal features can greatly increase the accuracy of models that predict pandemic trends.

**Figure 9.**
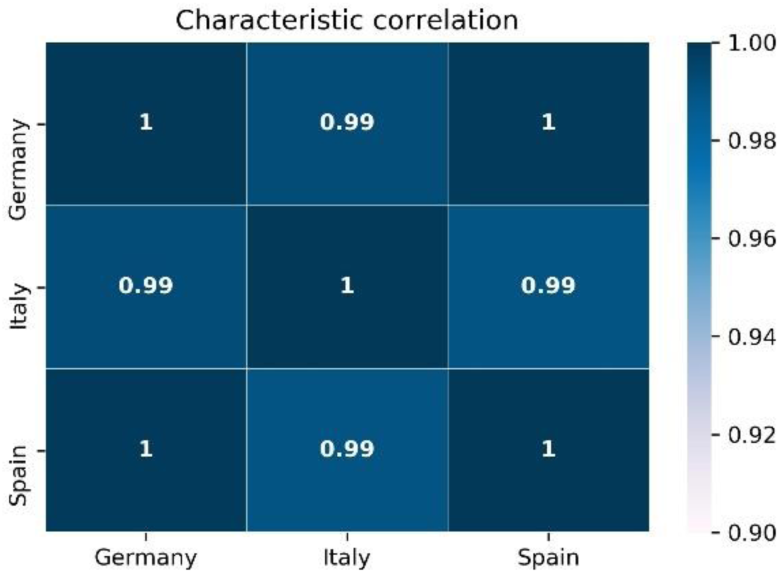
Correlation heatmaps of the accumulated number of confirmed cases in Germany, Italy, and Spain.

This study used a CNN, a GRU, a CNN-GRU, and COVID-19Net to predict the accumulated number of confirmed cases in Germany, Italy, and Spain (Fig. 10). The results indicated that the predictions produced by CNN-GRU were the least accurate and could not reflect any features or patterns. The GRU revealed a highly unstable increasing trend; hence, it could not be used as a reliable reference. The prediction results produced by CNN showed superiority over CNN-GRU and GRU. The main factor of the three countries had obvious spatial characteristics in terms of geographic location and it was unreasonable to consider only the time factor. Overall, the prediction result produced by the proposed COVID-19Net model was more accurate than that of the other models. Therefore, it can serve as a reliable reference.

**Figure 10.**
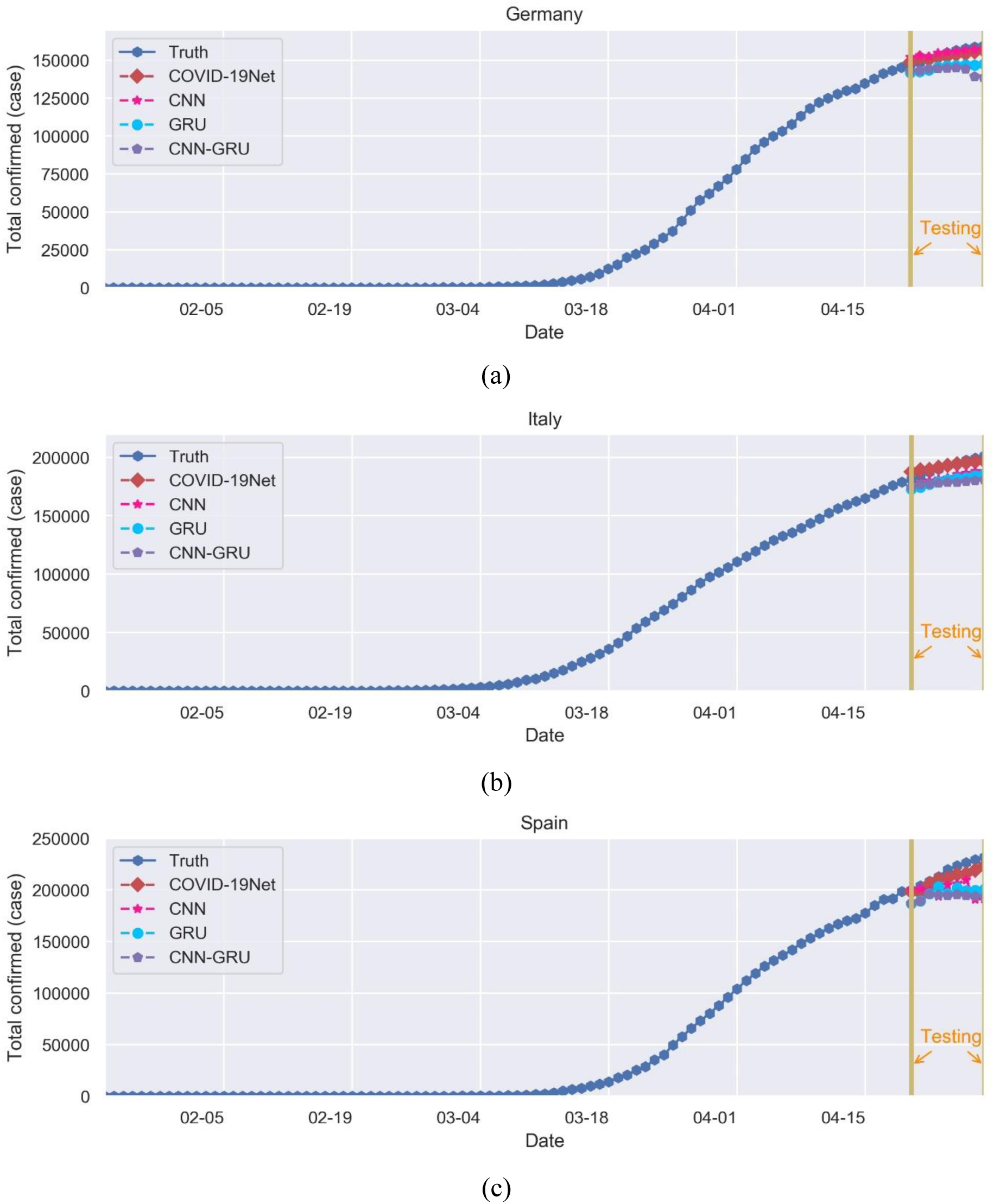
Results produced using the CNN, GRU, CNN-GRU, and COVID-19Net to predict the accumulated number of confirmed cases in (a) Germany, (b) Italy, and (c) Spain.

Tables I, II, and III list the MAE, MAPE, and RMSE values of the three countries obtained from the various models. Comparing these values verified that the proposed model was considerably more accurate than the other models and exhibited an MAPE value greater than 3 for the three countries. This indicated that COVID-19Net can accurately predict the accumulated number of confirmed cases in each country.

**Table 1.**
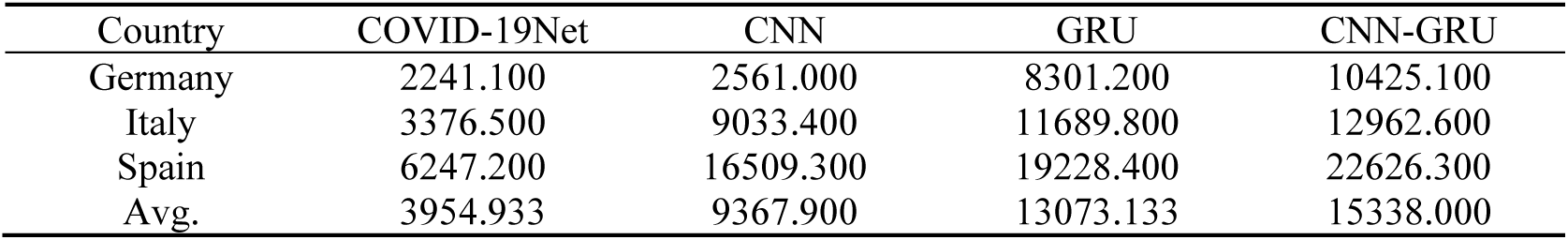
MAES OF THE THREE COUNTRIES.

**Table 2.** MAPES OF THE THREE COUNTRIES.

| Country | COVID-19Net | CNN | GRU | CNN-GRU |
| --- | --- | --- | --- | --- |
| Germany | 1.447 | 1.706 | 5.364 | 6.697 |
| Italy | 1.801 | 4.691 | 6.080 | 6.693 |
| Spain | 2.828 | 7.385 | 8.761 | 10.276 |
| Avg. | 2.025 | 4.594 | 6.735 | 7.889 |

**Table 3.** RMSES OF THE THREE COUNTRIES.

| Country | COVID-19Net | CNN | GRU | CNN-GRU |
| --- | --- | --- | --- | --- |
| Germany | 2549.030 | 3179.580 | 8772.430 | 11941.700 |
| Italy | 4051.940 | 9381.490 | 11996.600 | 14035.600 |
| Spain | 7183.000 | 21118.800 | 20824.900 | 24770.300 |
| Avg. | 4594.657 | 11226.623 | 13864.643 | 16915.867 |

Fig. 11 presents the bar chars of the MAE, MAPE, and RMSE values generated from COVID-19 Net, CNN, GRU, and CNN-GRU for Germany, Italy, and Spain. Fig. 11(a), 11(b), and 11(c) respectively correspond to Tables I, II, and III. The results indicated that the CNN-GRU model was the least accurate, and the GRU and CNN exhibited less favorable performance than did COVID-19Net. The models in descending order of prediction accuracy were COVID-19Net, CNN, GRU, and CNN-GRU, which verified that the proposed algorithm was the most accurate.

**Figure 11.**
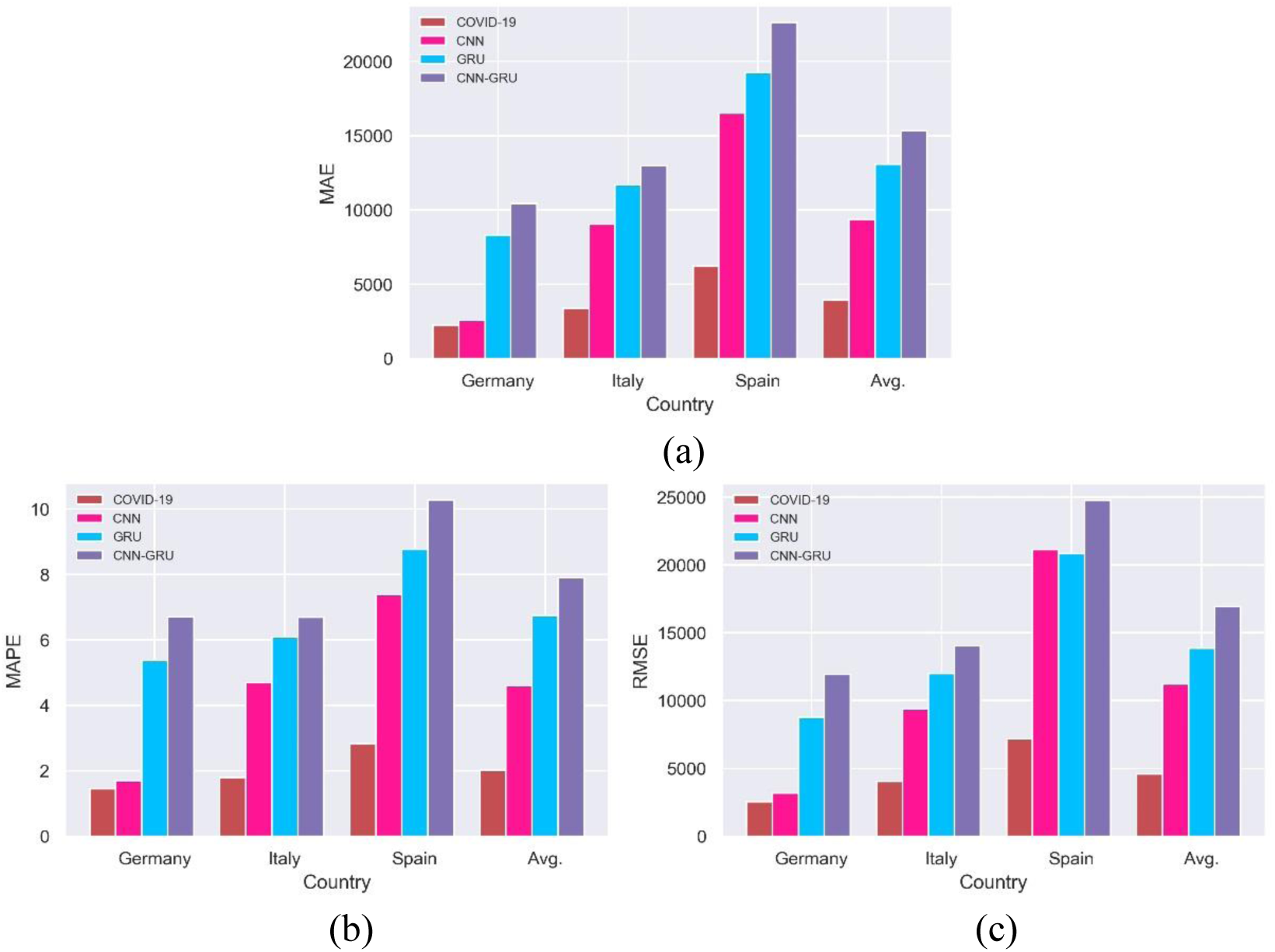
Bar charts of the (a) MAE, (b) MAPE, and (c) RMSE values derived from the prediction results of the three countries by using COVID-19Net, CNN, GRU, and CNN-GRU.

COVID-19 poses considerable challenges to healthcare systems worldwide. By using the proposed algorithm, we predicted that the demand for ward beds and ICU beds in Germany, Italy, and Spain would increase substantially, particularly before the pandemic peaks. If problems from lacking healthcare resources and social distancing cannot be resolved, demand will increase. COVID-19 may overwhelm the capacity of hospitals, particularly ICU nurses. The predicted values produced in this study can help countries develop and implement disease prevention measures and reduce gaps between the strategies employed by countries, including reducing services unrelated to COVID-19 prevention and temporarily increasing the capacity of the healthcare system. Based on the estimation results of Zhang et al. [17], It can be concluded that COVID-19 will end after the beginning of June, during which healthcare resources will be in heavy demand. However, this demand will also depend on social distancing measures implemented and other measures already imposed by each country. During this pandemic, relevant disease prevention measures must be maintained, and the importance of these measures must be highlighted to reduce the deaths of civilians and healthcare personnel.

## V. CONCLUSIONS

The likelihood of flattening the epidemic curve, as discussed in Western media, is overly optimistic because this entails no increase in COVID-19 cases. Currently, only China claims to have achieved this after implementing 2 months of lockdowns and strict measures. In this study, we proposed a novel model, COVID-19Net, to predict the accumulated number of confirmed cases in Germany, Italy, and Spain, which are heavily affected by the pandemic. The accumulated numbers of confirmed cases, deaths, and recovered cases and the daily numbers of newly confirmed cases, deaths, and recovered cases in the past 5 days were used to predict accumulated confirmed cases the next day. Comparing the prediction results and assessment indices of COVID-19Net, a CNN, a GRU, and a CNN-GRU verified that the CNN-GRU was the least accurate and generated an MAPE value of 1.447, 1.801 and 2.828 for the three countries. This indicated that CNNs cannot be used to extract features from the data of these countries. The hybrid model CNN-GRU also performed unfavorably. Although the CNN was slightly more accurate than the CNN and CNN-GRU, it remained unviable for predicting accumulated confirmed cases in the three countries. COVID-19Net was verified to be more accurate than the other three models because COVID-19 trend data contain spatiotemporal features that can be extracted using the deep neural network of COVID-19Net. The results of this study can serve as a crucial reference for devising public health strategies against COVID-19, and the proposed algorithm can serve as an effective tool for improving the allocation of hospital resources.

## Data Availability

Data were acquired from the daily situation reports disclosed by the WHO

## REFERENCES

1. Mahase, E., Covid-19: UK starts social distancing after new model points to 260 000 potential deaths. BMJ 2020, 368, m1089.

2. Dragomiretskiy, K.; Zosso, D., Variational Mode Decomposition. IEEE Transactions on Signal Processing 2014, 62, (3), 531–544.

3. Li, Q.; Feng, W.; Quan, Y. H., Trend and forecasting of the COVID-19 outbreak in China. J Infect 2020.

4. Nishiura, H.; Linton, N. M.; Akhmetzhanov, A. R., Serial interval of novel coronavirus (COVID-19) infections. Int J Infect Dis 2020.

5. Tang, B.; Bragazzi, N. L.; Li, Q.; Tang, S.; Xiao, Y.; Wu, J., An updated estimation of the risk of transmission of the novel coronavirus (2019-nCov). Infect Dis Model 2020, 5, 248–255.

6. Fan, C.; Liu, L.; Guo, W.; Yang, A.; Ye, C.; Jilili, M.; Ren, M.; Xu, P.; Long, H.; Wang, Y., Prediction of Epidemic Spread of the 2019 Novel Coronavirus Driven by Spring Festival Transportation in China: A Population-Based Study. Int J Environ Res Public Health 2020, 17, (5).

7. Roosa, K.; Lee, Y.; Luo, R.; Kirpich, A.; Rothenberg, R.; Hyman, J. M.; Yan, P.; Chowell, G., Real-time forecasts of the COVID-19 epidemic in China from February 5th to February 24th, 2020. Infect Dis Model 2020, 5, 256–263.

8. Zhong, L.; Mu, L.; Li, J.; Wang, J.; Yin, Z.; Liu, D., Early Prediction of the 2019 Novel Coronavirus Outbreak in the Mainland China Based on Simple Mathematical Model. IEEE Access 2020, 8, 51761–51769.

9. Nishiura, H., Backcalculating the Incidence of Infection with COVID-19 on the Diamond Princess. J Clin Med 2020, 9, (3).

10. Allam, Z.; Jones, D. S., On the Coronavirus (COVID-19) Outbreak and the Smart City Network: Universal Data Sharing Standards Coupled with Artificial Intelligence (AI) to Benefit Urban Health Monitoring and Management. Healthcare (Basel) 2020, 8, (1).

11. Yu, H.; Sun, X.; Solvang, W. D.; Zhao, X., Reverse Logistics Network Design for Effective Management of Medical Waste in Epidemic Outbreaks: Insights from the Coronavirus Disease 2019 (COVID-19) Outbreak in Wuhan (China). Int J Environ Res Public Health 2020, 17, (5).

12. McAleer, M., Prevention Is Better Than the Cure: Risk Management of COVID-19. Journal of Risk and Financial Management 2020, 13, (3).

13. Metsky, H. C.; Freije, C. A.; Kosoko-Thoroddsen, T.-S. F.; Sabeti, P. C.; Myhrvold, C., CRISPR-based surveillance for COVID-19 using genomically-comprehensive machine learning design. 2020.

14. Guo, Q.; Li, M.; Wang, C.; Wang, P.; Fang, Z.; tan, J.; Wu, S.; Xiao, Y.; Zhu, H., Host and infectivity prediction of Wuhan 2019 novel coronavirus using deep learning algorithm. 2020.

15. Yang, Z.; Zeng, Z.; Wang, K.; Wong, S.-S.; Liang, W.; Zanin, M.; Liu, P.; Cao, X.; Gao, Z.; Mai, Z.; Liang, J.; Liu, X.; Li, S.; Li, Y.; Ye, F.; Guan, W.; Yang, Y.; Li, F.; Luo, S.; Xie, Y.; Liu, B.; Wang, Z.; Zhang, S.; Wang, Y.; Zhong, N.; He, J., Modified SEIR and AI prediction of the epidemics trend of COVID-19 in China under public health interventions. Journal of Thoracic Disease 2020, 12, (3), 165–174.

16. Anastassopoulou, C.; Russo, L.; Tsakris, A.; Siettos, C., Data-Based Analysis, Modelling and Forecasting of the COVID-19 outbreak. 2020.

17. Zhang, X.; Ma, R.; Wang, L., Predicting turning point, duration and attack rate of COVID-19 outbreaks in major Western countries. Chaos Solitons Fractals 2020, 109829.

18. Kucharski, A. J.; Russell, T. W.; Diamond, C.; Liu, Y.; Edmunds, J.; Funk, S.; Eggo, R. M.; Sun, F.; Jit, M.; Munday, J. D.; Davies, N.; Gimma, A.; van Zandvoort, K.; Gibbs, H.; Hellewell, J.; Jarvis, C. I.; Clifford, S.; Quilty, B. J.; Bosse, N. I.; Abbott, S.; Klepac, P.; Flasche, S., Early dynamics of transmission and control of COVID-19: a mathematical modelling study. The Lancet Infectious Diseases 2020.

19. Huang, C.-J.; Chen, Y.-H.; Ma, Y.; Kuo, P.-H., Multiple-Input Deep Convolutional Neural Network Model for COVID-19 Forecasting in China. 2020.

20. WHO Situation Reports:https://www.who.int/emergencies/diseases/novelcoronavirus-2019/situation-reports.

21. Github:https://ctc.github.io/COVID-19-europe.

22. Kuo, P.-H.; Huang, C.-J., An Electricity Price Forecasting Model by Hybrid Structured Deep Neural Networks. Sustainability 2018, 10, (4).

23. Wang, Y.; Chen, Q.; Ding, M.; Li, J., High Precision Dimensional Measurement with Convolutional Neural Network and Bi-Directional Long Short-Term Memory (LSTM). Sensors (Basel) 2019, 19, (23).

24. Hsieh, T.-H.; Kiang, J.-F., Comparison of CNN Algorithms on Hyperspectral Image Classification in Agricultural Lands. Sensors 2020, 20, (6).

25. Zhang, M.; Geng, G.; Chen, J., Semi-Supervised Bidirectional Long Short-Term Memory and Conditional Random Fields Model for Named-Entity Recognition Using Embeddings from Language Models Representations. Entropy 2020, 22, (2).

26. Chen; Xie; Yuan; Huang; Li, Research on a Real-Time Monitoring Method for the Wear State of a Tool Based on a Convolutional Bidirectional LSTM Model. Symmetry 2019, 11, (10).

27. Lin, H.; Sun, Q.; Chen, S.-Q., Reducing Exchange Rate Risks in International Trade: A Hybrid Forecasting Approach of CEEMDAN and Multilayer LSTM. Sustainability 2020, 12, (6).

28. Gu, Z.; Li, Z.; Di, X.; Shi, R., An LSTM-Based Autonomous Driving Model Using a Waymo Open Dataset. Applied Sciences 2020, 10, (6).

29. Batur DİNler, Ö.; Aydin, N., An Optimal Feature Parameter Set Based on Gated Recurrent Unit Recurrent Neural Networks for Speech Segment Detection. Applied Sciences 2020, 10, (4).

30. Dutta, A.; Kumar, S.; Basu, M., A Gated Recurrent Unit Approach to Bitcoin Price Prediction. Journal of Risk and Financial Management 2020, 13, (2).

31. Jaihuni, M.; Basak, J. K.; Khan, F.; Okyere, F. G.; Arulmozhi, E.; Bhujel, A.; Park, J.; Hyun, L. D.; Kim, H. T., A Partially Amended Hybrid Bi-GRU—ARIMA Model (PAHM) for Predicting Solar Irradiance in Short and Very-Short Terms. Energies 2020, 13, (2).

32. Cho, H.; Yoon, S. M., Divide and Conquer-Based 1D CNN Human Activity Recognition Using Test Data Sharpening. Sensors (Basel) 2018, 18, (4).

33. Zhang, E.; Xue, B.; Cao, F.; Duan, J.; Lin, G.; Lei, Y., Fusion of 2D CNN and 3D DenseNet for Dynamic Gesture Recognition. Electronics 2019, 8, (12).

